# A Modelling Analysis of Strategies for Relaxing COVID-19 Social Distancing

**DOI:** 10.1101/2020.05.19.20107425

**Authors:** George J Milne, Simon Xie, Dana Poklepovich

## Abstract

**Background:** The ability of countries to contain and control COVID-19 virus transmission via social distancing is critical in the absence of a vaccine. Early activation of robust measures has been shown to control the daily infection rate, and consequential pressure on the health care system. As countries begin to control COVID-19 spread an understanding of how to ease social distancing measures to prevent a rebound in cases and deaths is required.

**Methods:** Using COVID-19 transmission data from the outbreak source in Hubei Province, China prior to activation of containment measures, we adapted an established individual-based simulation model of the city of Newcastle, Australia. Simulation of virus transmission in this model, with and without, social distancing measures activated permitted us to quantify social distancing effectiveness. Optimal strategies for relaxing social distancing were determined under two settings: with high numbers of daily cases, as in New York; and where early social distancing activation resulted in limited ongoing transmission, as in Perth, Australia.

**Findings:** In countries where strong social distancing measures were activated after the COVID-19 virus had spread widely, our study found these measures are required to be maintained for significant periods before being eased, to return to a situation where daily case numbers become low. In countries where early responses to the COVID-19 pandemic have been highly successful, as in Australia, we show that a staged relaxation of social distancing prevents a rebound in cases.

**Interpretation:** Modelling studies and direct observation have shown that robust and timely social distancing have the most effect in containing the spread of the COVID-19 virus. Questions arise as to the duration of strong social distancing measures, given they are highly disruptive to society and economic activity. This study demonstrates the necessity of holding robust social distancing in place until COVID-19 virus transmission has significantly decreased, and how they may then be safely eased.

## Introduction

At the early stages of the COVID-19 coronavirus pandemic first detected in Wuhan, China data on virus transmissibility and pathogenesis was uncertain, as would be the case for a novel influenza virus.^1^ Given this uncertainty, Chinese authorities adopted strict measures to contain COVID-19 spread, by continuing closure of schools and workplaces, which were already closed for the Chinese Lunar New Year, activating measures to enforce significant community contact reduction, and closing transport links between population centres. This response was in contrast to the situation which occurred in 2002/3 with the Severe Acute Respiratory Syndrome epidemic, where the SARS coronavirus had spread widely before measures were effected to contain it.^2^

We used early transmission data from the unmitigated COVID-19 outbreak in China,^3^ to adjust parameters in an established, individual-based simulation model of an Australian city to reflect these COVID-19 characteristics, and applied the model to evaluate the effectiveness of a range of social distancing interventions. The results provide guidance to public health authorites as to the mitigating effect of individual social distancing measures, such as school and workplace closure and reductions in community-wide contact, and under alternative levels of “robustness”.^4^ Similar modelling studies have also evaluated social distancing in mitigating COVID-19 virus spread.^5-9^

Public Health authorities also require guidance as to how best to relax social distancing measures to allow an economy to restart while preventing a “bounce back” second peak in case numbers. We thus evaluated how social distancing measures may be relaxed in an optimal manner that achieves the following two goals: to allow the economy to grow again, from a low activity base affected by workplace closures; and to do this in a way that prevents a rebound in case numbers and deaths. To determining the impact of relaxing social distancing we conducted analyses under two distinct settings: New York City, a high transmission setting where an exponential growth in cases occurred prior to social distancing activation on 22^nd^ March 2020, with a strengthening of measures occurringon 12^th^ April 2020;^10^ and a low transmission setting of Perth, Australia where strict border closure and social distancing interventions were activated prior to significant within country COVID-19 virus transmission.^11^

In Australia robust social distancing measures were adopted country-wide starting on 20^th^ March 2020 and further strengthened on 23^rd^ and 26^th^ March 2020,^11-13^ resulting in significant reductions in workplace and community-wide contact. These measures resulted in very low levels of COVID-19 virus transmission, with almost all diagnosed cases being returing Australians infected outside the country, or those who had been in direct contact with such individuals. In Western Australia, closure of State borders coupled with rigorous contact tracing and testing prevented any significant outbreak occurring in metropolitan Perth, a conurbation with a population of approximately 2·2 million.

Under a New York City scenario we evaluated the timing of social distancing relaxation, to determine the earliest period when they may be eased without such changes causing an increase in the growth of infections. For the Perth, Australia scenario we analysed a staged relaxation of social distancing, starting with schools reopening after the Easter holidays, then an increase in the limit of gatherings from two persons to ten, and the opening of cafes and restaurants and increased return to the workplace. We further analysed a scenario where there was a rapid increase in cases due to introduction of a number of asymptomatic individuals, such as may occur when borders are reopened.

The results directly address questions related to the timing and scale of social distancing relaxation measures required, with the aim of increasing economic activity while preventing a rebound in virus transmission.

## Methods

A community-based simulation model capturing the demographics and movement patterns of individuals in a city together with the virus transmission characteristics of COVID-19, was used to evaluate the effectiveness of a range of social distancing strategies. The model is individual-based (*c.f*. agent-based) and represent each individual in a specific community, matching recent census and other government data.^14,15^ Related individual-based simulation models for multiple population centres in Australia, and in South Africa, Thailand, Vietnam and Papua New Guinea, have been previously developed using the same underlying automata-theoretic modelling methodology to capture the dynamics of both pathogen transmission and population mobility.^16-20^

This approach to disease modelling has allowed us to explicitly simulate person-to-person virus transmission, the probability of such transmission, the location of transmission (*e.g*. school, workplace, home, community) and determines each individual’s infection status through time.^17-19,21^ Such simulation models create a “virtual world” of individuals whose daily movement, changing contact patterns and disease biology dynamics aim to replicate that of the real-world system in as much detail as data sources permit, such as data from the POLYMOD contact pattern study.^22^

As with the SARS outbreak in 2003 no vaccine is available,^23^ and the population has no immunity at the outset. Reliance must be made on robust social distancing interventions, contact tracing and early isolation of diagnosed cases. The aim of social distancing interventions in the current COVID-19 situation is to slow down transmission and reduce the growth rate in case numbers. This approach aims to lessen the daily pressure on health care personnel and hospital facilities, such as intensive care beds, and to lower mortality rates.

This study used an established, individual-based model of the city of Newcastle, New South Wales, Australia. This model matches the real-world counterpart with respect to population size, household structure, age of individuals in each household (stratified from Australian census data into ten age bands), employment, schooling, and daily movement between these locations. The model was developed using detailed census, workplace and mobility data using a model development methodology applied previously.^18^ Such models create realistic representations of the respective communities at an individual-by-individual level and aim to use the best available data sources, including from the Australian Bureau of Statistics (ABS). ^24^

The Newcastle model represents 272,407 people in an urban area. ABS census data^14,15^ were used to capture the age-specific demographics of every household in the community. Data for schools, including geographical location and pre-primary, primary and secondary enrolment numbers for each school in the Newcastle were obtained from New South Wales state government publications.^25^ ABS data were also used to determine workplace locations and workforce sizes. This data was used to generate a model which captures the movement and contact patterns of individuals on a day-by-day basis.^22^

Simulation model parameter settings to reflect the transmission characteristics of the COVID-19 epidemic were determined via calibration to represent an unmitigated outbreak with a basic reproduction number R_0_ of approximately 2·2, taken from work by Li and colleagues.^3^ This basic reproduction number corresponds to that derived by Kucharski and colleagues.^26^ This data provided virus transmission settings for the model corresponding to the spread characteristics in Wuhan, China prior to activation of social distancing measures on 23^rd^ January 2020. These parameter settings provided an unmitigated epidemic baseline with which to compare alternative social distancing (SD) strategies. Model outputs obtained by running the simulation software for the duration of an infectious disease outbreak gave the infection history of each individual in the modelled community. This data was used to determine the total number of infectious individuals, where and when infection occurs, and may thus be used to quantify the resulting health burden, *i.e*. hospitalisations and deaths.^21,27^

To determine the effectiveness of specific social distancing interventions we compared the number of daily infections generated by an unmitigated COVID-19 outbreak with one which has specific social distancing measures in place. Modelling experiments were conducted for alternative social distancing strategies; the difference in cases quantifying intervention effectiveness.

A transmission parameter is used to model the probability of COVID-19 transmission following contact between a susceptible and an infectious individual. That is, a pairing of an individual in infectious state *I* and one in susceptible state *S*, as in the *S-E-I-R* state transition representation of the spread dynamics of a virus.^28^ Adjusting this transmission parameter allowed us to replicated epidemics with different reproduction numbers, and thus attack rates.

Assumed transmission characteristics of COVID-19 ^3^ are as follows: a basic reproduction number R_0_ of 2·2; an incubation period averaging 5·5 days, from infection to symptom emergence (if any); a latent period averaging 4·5 days, from infection to becoming infectious; an infectious period averaging 3·0 days, the first day being asymptomatic; and a 35% asymptomatic rate, that is, 35% of infectious individuals show no symptoms and are not classed as cases. These data are contained in Table 1. At the early stage of the COVID-19 epidemic reliable age-specific symptomatic attack rates were unavailable and we thus assumed transmission between different ages of individuals occurs similarly.

**Table 1:** Summary of simulation parameters.

| Parameter | Value |
| --- | --- |
| Asymptomatic Rate | 35% |
| Targeted $R_0$ | 2.2 |
| Simulation Output Infection Rate | 66.00% |
| Infection Related |  |
| Latent Period (Exposed) | 4.5 days |
| Incubation Period (Including Latent) | 5.5 days |
| Infectious Period | two days (Finish at day 7.5) |
| Post-symptomatic Period | None |
| Symptomatic Case Diagnosis Rate | 50% |
| (Basic) Case Isolation | 50% adults, 90% children |

The model explicitly represents each household, workplace and school in a community and the movement of individuals between, as they move from households to schools and workplaces in a daytime cycle, then return to their household in an evening cycle, with each day split into a day and night period. This mobility mechanism allowed us to model changing contact patterns, with virus transmission occurring in these contact locations, and also in the wider community, including at weekends. The model represents the individual-to-individual contact patterns in as much detail as data sources provide, to accurately describe how the movement of individuals allows virus transmission to spread over a geographic region. This level of detail is critical for modelling social distancing interventions, whose aim is to minimize person-to-person contact patterns and consequential virus transmission.

### Social distancing

Four distinct social distancing measures are available to health authorities: *a*) school closure; *b*) workplace closure and non-attendance; *b*) symptomatic case isolation; *d*) reduced community-wide contact. Assumptions regarding feasible social distancing measures made in prior pandemic influenza modelling studies are also applicable in the novel coronavirus context.^2,3^ We considered both the strength and timing of social distancing activation, allowing us to analyse modifying social distancing as the dynamics of a localised epidemic changes through time.

Social distancing measures are briefly explained as follows. School closure: when schools and further education institutions are closed students have contact with household members during the daytime and also have contact in the community. Workplace non-attendance and thus contact reduction: a percentage of all persons in the workforce are absent, have contact within their household in the daytime and still having contact in the wider community. Increased case isolation: 90% of adults and 100% of children withdraw to the home on becoming ill *i.e*. are symptomatic cases and only have contact with household members, an increase from a baseline assumption of 50% adults and 90% children who withdraw when symptomatic. Community contact reduction: contact in the wider community is reduced by a given percentage to reflect the strength of this intervention.

### Relaxing social distancing

There is significant interest in when and how to relax social distancing interventions once daily case numbers have declined to a level which is sustainable by available health care facilities, in terms of ICU bed and staffing resources. To analyse relaxation strategies we considered two distinct situations, *a*) where daily diagnosed case numbers are high and either increasing or leveling out, and *b*) where case numbers are very low, and a localised epidemic is tending to elimination. In this latter situation we analysed response strategies needed for a rapid increase in case numbers, such as may occur when borders are reopened.

### Role of the funding source

The Department of Health, Government of Western Australia provided guidance on certain social distancing measures included in the study. They had no further role in the study design, the analysis and interpretation of data, nor in writing the report and the decision to submit for publication.

## Results

### Relaxing Social Distancing for a High Transmission Setting

Results are presented for a high COVID-19 transmission setting, using New York City as a representative example. Initial diagnosed case numbers for New York State were used to fit the model to the unmitigated, early stage epidemic which sustained an exponential growth rate, as in Figure 1, blue curve. The New York State activated social distancing measures on 22^nd^ March 2020, which arrested the epidemic growth rate, as in the orange curve in Figure 1. This was followed by a strengthening of social distancing measures on 22^nd^ April 2020, when we estimate workplace nonattendance (WN) and community-wide contact reduction (CCR) increased from 35% to 50%. This strengthening of social distancing is predicted to significantly reduce the daily infection rate, as in the grey curve, and subsequent daily case and mortality rates, by reducing high virus transmission rates.

**Figure 1:**
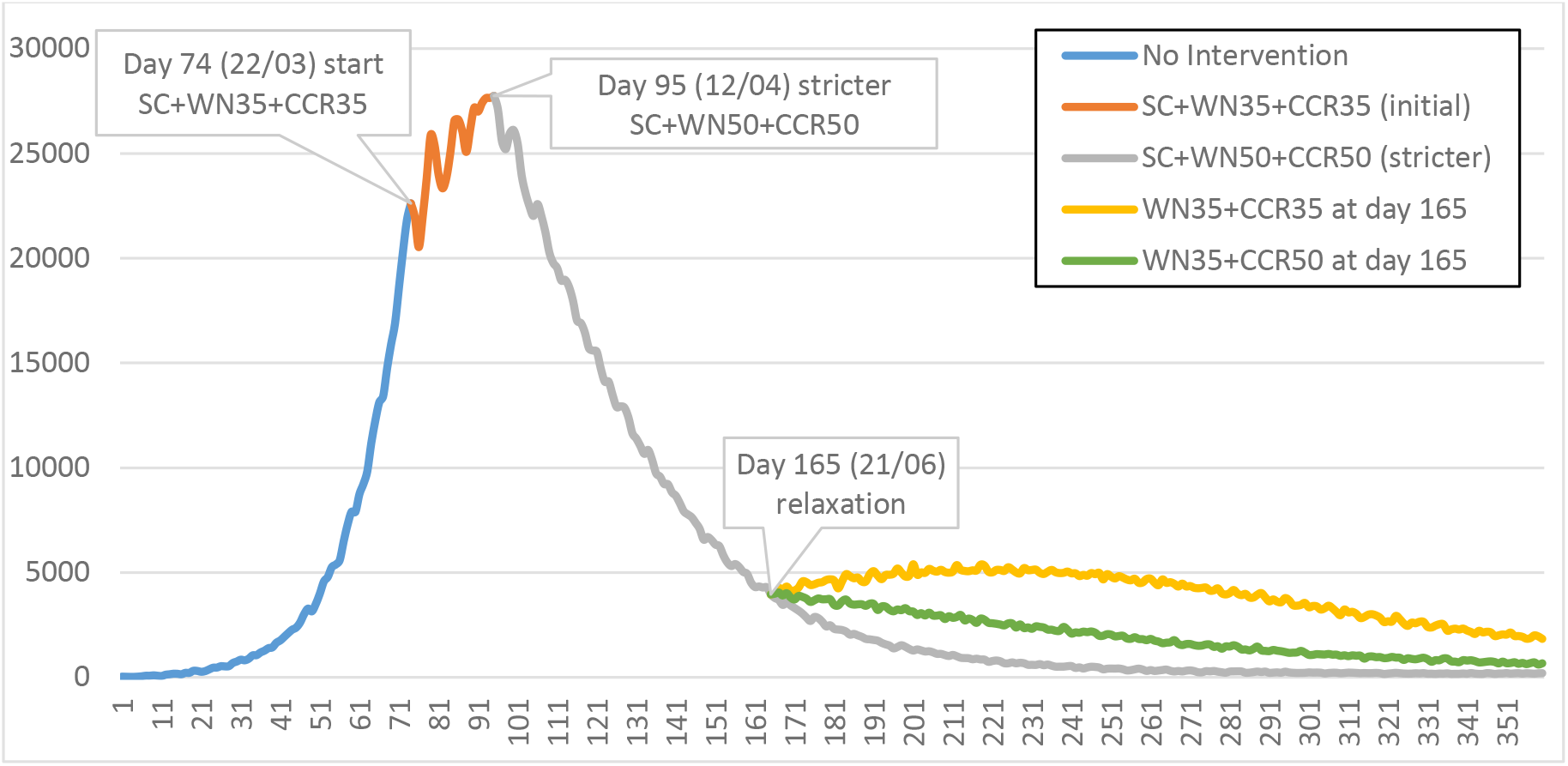
epidemic curve for New York City, population 8·4 million, with social distancing activation and easing. Initial unmitigated epidemic coloured blue. School closure and estimated 35% reduction in workplace and community-wide contact activated 22nd March 2020 given by orange curve. Social distancing strengthened 12th April 2020 by increased reduction in workplace and community contact to 50%, schools remained closed, grey curve. Schools reopen 21st June 2020 with social distancing interventions maintained, illustrated by green curve. School reopening together with workforce and community-contact increased to 65% given by orange curve. All social distancing strategies assume case isolation of 90% children and 50% adults.

Figure 1 presents daily *infection* data, which may be transformed into a lower diagnosed case rate and subsequent case fatality rate. The infection rate is seen to be immediately impacted by activation of social distancing measures, as transmission rates are quickly reduced by the reduction in person-to-person contact. The impact of social distancing measures activated in NYC in March and April would have been detected by reductions in diagnosed cases approximately seven to ten days later, due to the incubation period delay followed by delays in diagnosis. There would be a similar delay in the reduction in deaths due to the introduction and further strengthening of the social distancing measures.

Figure 1 illustrates two scenarios where social distancing interventions are relaxed in a manner which permits an economic “restart” while keeping the infection rate at a level that may be managed by available health care reources. Here we assume schools reopen on 21^st^ June 2020, allowing parents providing childcare of young children to return to the workplace. We evaluated school reopening with social distancing interventions maintained, as illustrated by the green curve in Figure 1. An alternative scenario is when schools restart simultaneously with workplace attendance increasing from 50% to 70% (workplace non-attendance WN reduced to 30%) and community-contact increasing from 50% to 65% (community-contact reduction reduced to 35%).

Activation of strong social distancing measures are seen to result in a declining rate of daily infections, the grey curve. If schools reopen on 21^st^ June 2020, when the New York City infection rate is predicted to be of the order of 4,000 new infections per day, the green epidemic curve indicates that this reduces the rate of decline in infections. However it also shows that the daily infection rate is still declining, though at a slower rate.

If school reopening is coupled with a simultaneous increase in workforce attendance (to 65%) and increased community-wide contact (to 65%), as with the orange curve, the decline in daily cases is arrested and the epidemic curve rebounds. This rebound increases slowly up to 1,000 additional infections per day, with this rebound peak occurring approximately two months later. Assuming a 35% asymptomatic rate and a 50% diagnosis rate for symptomatic infections, this 1,000 additional infections translates into approximately 300 additional diagnosed cases per day, a number that may be managed by available health care resources.

Figure 2 presents results for alternative social distancing relaxation scenarios, where the effect of the timing of reduction in strength of measures is determined. We considered the same combined reduction as illustrated in Figure 1, involving reopening of schools, and increasing workplace attendance and community-wide contact, from 50% to 65%. The timing of this relaxation of social distancing was evaluated at six, eight, ten, twelve and fourteen weeks after the measures were strengthened on 12 April 2020.

**Figure 2:**
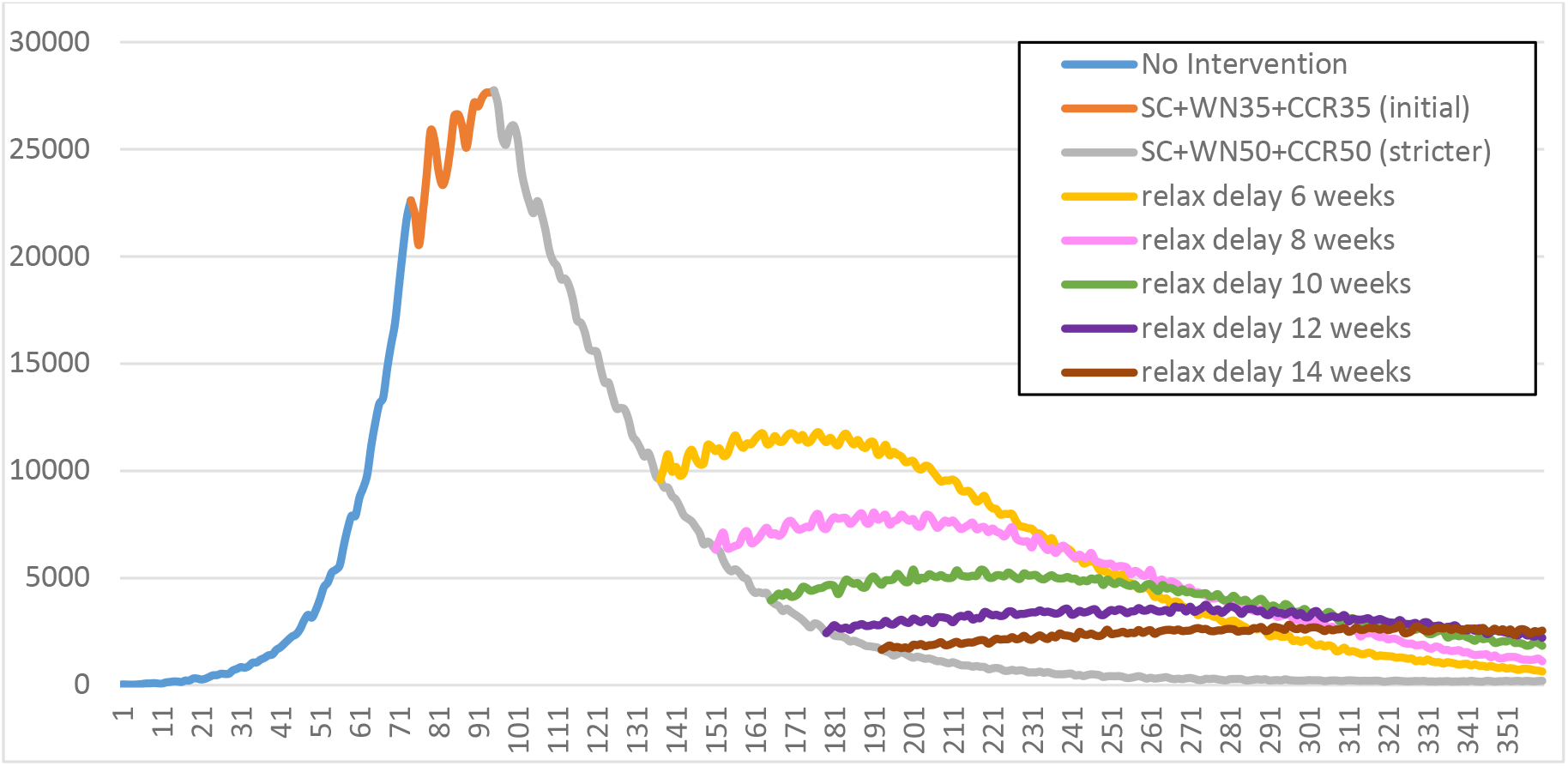
timing the introduction of weaker social distancing measures.

It is apparent that the earlier the robust social distancing measures are relaxed the greater the period of sustained high transmission. The six (yellow) and eight week (pink) relaxation curves illustrate that the rebound in daily infections is moderate and there is a significant period of where the infection rate is high, between 7,500 and 12,000 new infections each day. This situation contrasts with a continuous decline in infections if strong social distancing is maintained continuously, the grey curve. Holding the strong social distancing in place for 12 or 14 weeks before relaxation gives a flat plateau effect in the daily infection rate, the purple and brown curves. These occur on 8^th^ and 23^rd^ July respectively, with sustained infection rates between 2,000 and 3,000 per day.

For the six epidemic curves in Figure 2, the cumulative number of infections occurring between 23^rd^ May 2020 (day 137) and 29^th^ August 2020 (day 236) are as in Table 2.

**Table 2:** infections from day 137 to day 236 with alternative relaxation timing; social distancing changing from SC+WN50+CCR50 to WN35+CCR35.

| Timing of relaxation | Infections from day 137 to day 236 |
| --- | --- |
| 6 weeks | 1,017,755 |
| 8 weeks | 739,402 |
| 10 weeks | 532,254 |
| 12 weeks | 405,716 |
| 14 weeks | 345,252 |
| No relaxation | 307,786 |

### Relaxing Social Distancing in a Low Transmission Setting

In Western Australia, starting on 15^th^ March 2020, large gatherings were banned, restaurants, entertainment and sporting venues were closed, people were encouraged to work from home if possible, and restrictions on contact outside of households was restricted. Schools were not required to close, though some independent schools did close, and across all schools parents were able to keep children at home if they wished, with home and on-line schooling occurring. Universities stopped in-person teaching with on-line lectures streamed to students.

These measures, coupled with rigorous contact tracing, testing and quarantining of diagnosed cases resulted in low levels of COVID-19 virus transmission. This has prevented any significant COVID-19 outbreak in Perth, a conurbation with a population of approximately 2·2 million. With state and international borders closed, and returing Australians being quarantined for 14 days in hotels, the COVID-19 virus in Western Australia is tending to elimination status.

To inform the Government of Western Australia on strategies for the safe easing of social distancing we modelled a staged relaxation of physical distancing measures, starting with schools reopening after the Easter holidays, then an increase in the limit of gatherings from two persons to ten, then the opening of cafes and restaurants, and increased numbers returning to the workplace. We further analysed a scenario where there was a rapid increase in cases, with a spike in infections being a possibility once borders reopen.

While school closure did not occur in Western Australia, parents were encouraged to keep children at home if possible. The social distancing measures activated in mid-March were modelled as follows: school and workplace attendance was reduced to 50% (SC50 and WN50), community-wide contact was reduced by 80% (CCR80) and increased case isolation (ICI) occurred, with 100% of children and 90% of adults isolated on case diagnosis.

The effect on the daily infection rate in Perth following activation of such robust social distancing measures on 15^th^ March 2020, denoted by (SC50+ICI+ WN50+CCR80), is illustrated by the orange curve in Figure 3. The initial unmitigated growth in infections (blue) has its increasing infection growth rate arrested by these robust social distancing measures.

**Figure 3:**
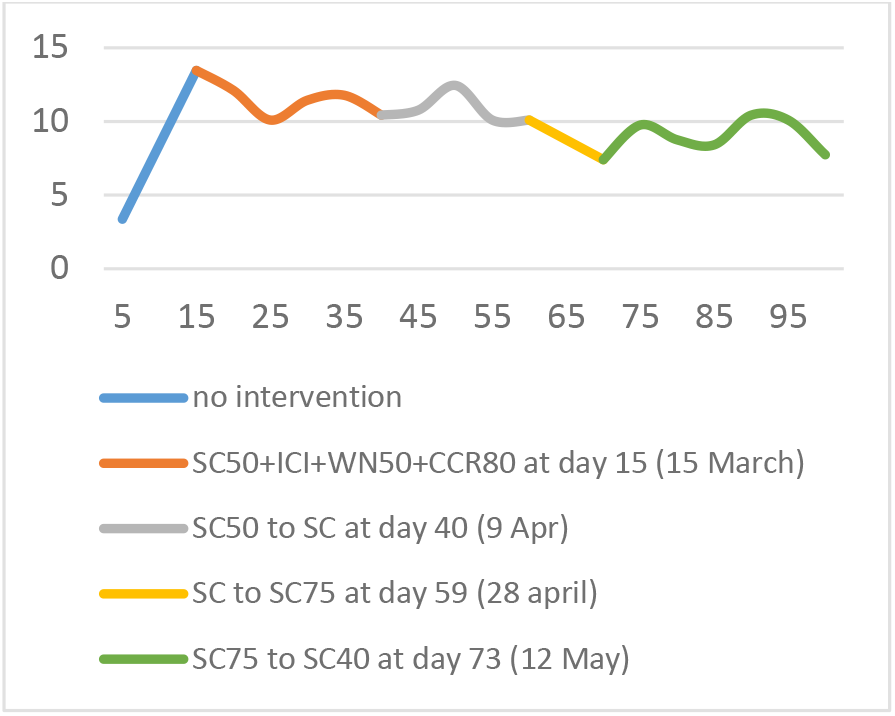
school closure and partial reopening.

The Easter school holidays occurred on 9^th^ April 2020 with all schools closing, and the impact of this slight change to the strong measures already in place is illustrated by the grey curve, with a slightly reducing trend in infections. The yellow and green predicted daily infections in Figure 3 model the effect of a staged reopening of schools, while holding all other social distancing measures in place. These involve 25% of all students (SC75) returning to school on 28^th^ April 2020 and 60% attending (SC40) from 12^th^ May 2020 onwards.

Given the low incidence of COVID-19 virus transmission and the stochastic nature of contact patterns captured in the model, infection occurrence as illustrated in Figure 3 averaged between five and 12 infections per day for the 2·2 million population of Perth. Given a 35% asymptomatic rate and assuming a 50% diagnosis of symptomatic cases (see Table 1) this resulted in between 1·6 and four diagnosed cases per day, with only a slight variation being due to the time-varying school attendance rates. This rate of diagnosed cases corresponds closely to the situation in Perth up to 20^th^ April 2020.^29^

To determine the impact of relaxing social distancing, with the aim of increasing workplace participation and thus increasing economic activity, we considered three alternative social distancing relaxation strategies to take effect on 1^st^ June 2020, as in Figure 4. Workplace contact restrictions were all reduced to 20% nonattendance (WN20), allowing 80% of the workforce to return to pre-pandemic levels. Alternative community-wide contact reductions evaluated were changes from 80% to 65%, 50% and 40% and are denoted by CCR65 (brown), CCR50 (purple) and CCR40 (pink) respectively in Figure 4.

**Figure 4:**
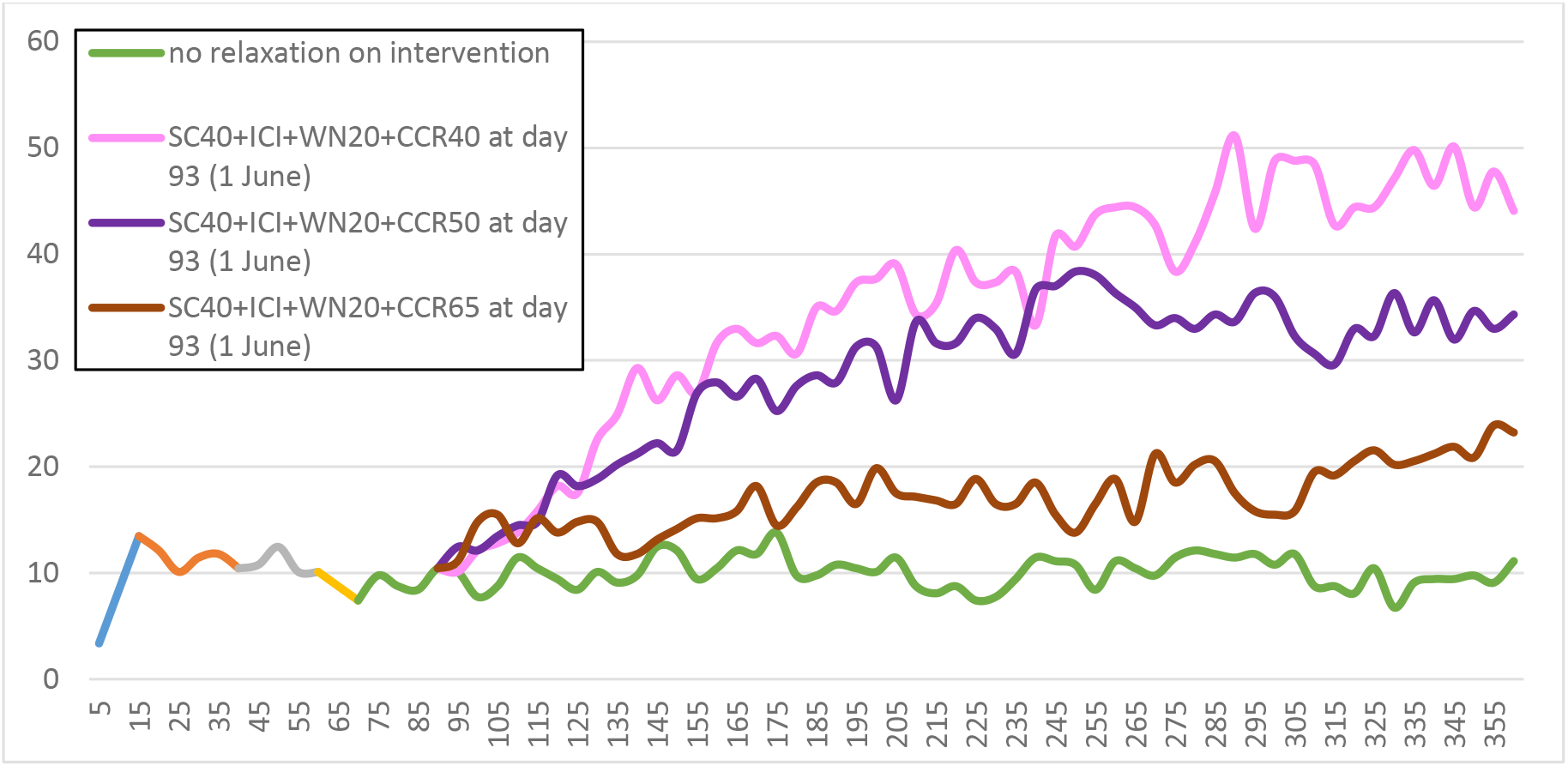
relaxing workplace and community-wide contact restrictions.

The strong social distancing measures in effect prior to 1^st^ June 2020, namely SC40+ICI+WN50+CCR80, were adjusted to the following settings: *a)* SC40+ICI+WN20+CCR40 (pink curve); *b)* SC40+ICI+WN20+CCR50 (purple curve); and to *c)* SC40+ICI+WN20+CCR65 (brown curve), with results of simulations illustrated in Figure 4. If social distancing is held at pre-June levels, the green graph indicates a steady number of approximately ten new infections each day, resulting in approximately three diagnosed cases per day, from assumptions in Table 1.

A substantial increase in workplace activity due to WN50 changing to WN20, coupled with a slight increase in community contact (CCR80 to CCR65) results in the gently increasing brown infection curve, averaging 16 infections and six diagnosed cases over the nine month period modelled. If the increase in workplace participation is coupled with an increase in community-wide contact to CCR50, the infection rate increases to an average of 34 per day, with approximately ten diagnosed cases. When the increase in workplace participation is coupled with a higher rate of community-wide contact, of CCR40, the infection rate increases steadily reaching a peak of approximately 50 daily infections, and may result in 17 diagnosed cases.The key difference between scenario *a* and scenarios *b* and *c* is the latter two settings have lower community-wide contact reductions, 50% and 65% respectively, compared to 40%.

It should be noted that the simulation experiments illustrated in Figure 3 have two infectious individuals seeded into the population of Perth each day. While Western Australia is approaching COVID-19 virus elimination, as of 30^th^ April 2020, this seeding prevents the daily infection rate from falling to zero.

Australia introduced strict international border controls on 20 March 2020, only permiting Australian residents to return and requiring that they were quarantined for 14 days.^30^ Similarly, the borders of Western Australia were also closed. To analyse a situation which may arise when interstate or international borders reopen, we evaluated a scenario where a number of asymptomatic infectious persons arrive in Perth and are not quarantined. This assumed the same changes to social distancing as occurred on 1^st^ June 2020 in the previous analyses, but with the introduction of 30 infectious persons on 1^St^ July 2020. The daily changes in infections under the four social distancing measures are illustrated in Figure 5.

**Figure 5:**
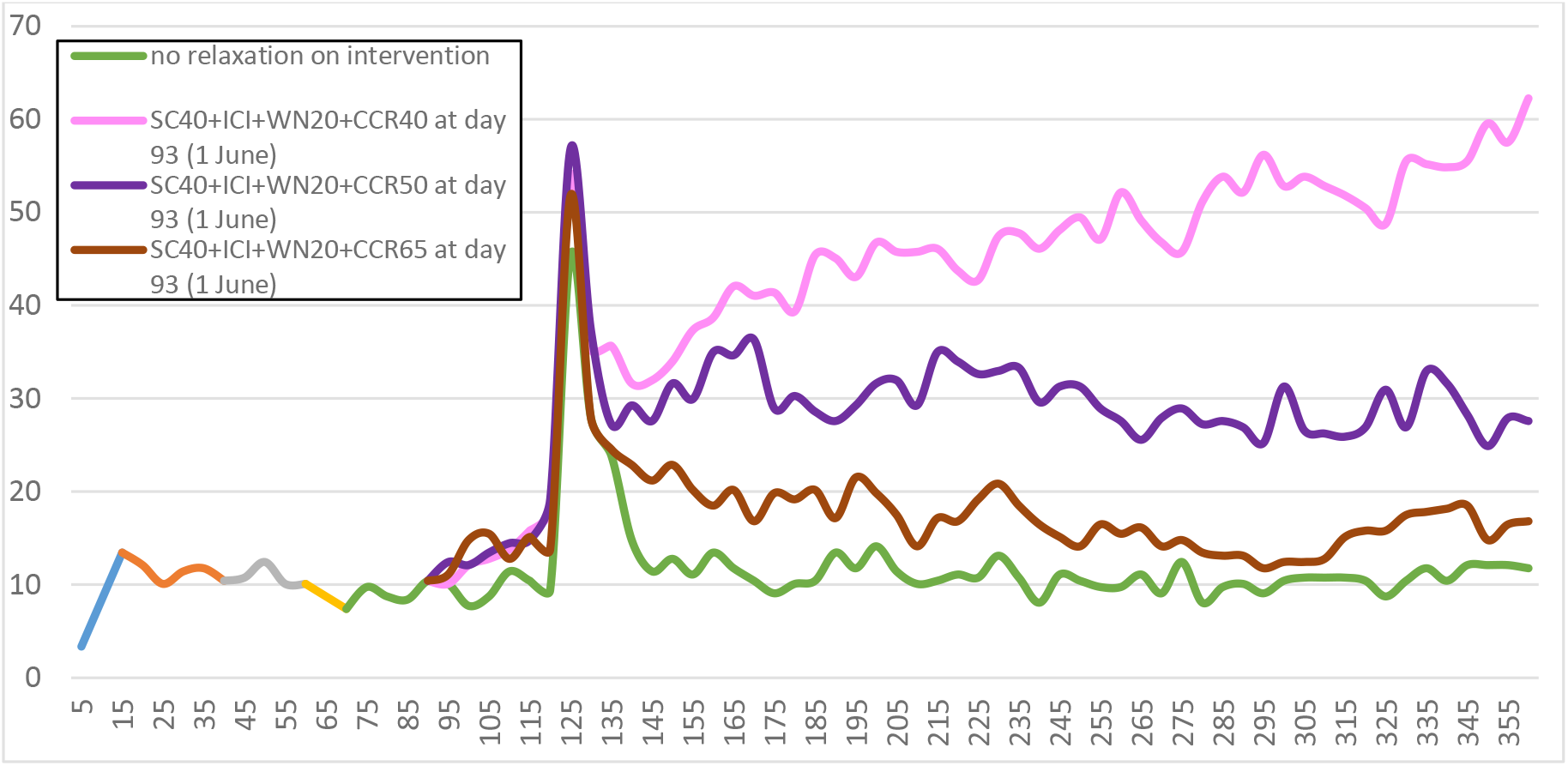
surge in infections due to border reopening.

From Figure 5 the rapid increase in infections due to the arrival of 30 infectious individuals into metropolitan Perth, a population of approximately 2·2 million, has little long-term effect when compared to the setting illustrated in Figure 4. The sudden simultaneous introduction of cases into the community is seen to have no lasting effect on the daily infection rate after approximately three weeks.

The largest relaxation of social distancing, to SC40+ICI+WN20+CCR40 which results in the pink curves in Figures 4 and 5, increases the infection rate to a peak of approximately 50 and 60 daily infections respectively. The two remaining changes to social distancing, where community contact reductions are eased to SC40+ICI+WN20+CCR50 and SC40+ICI+WN20+CCR65, represented by the purple and brown graphs respectively, do differ slightly between Figure 4 and Figure 5. The purple graphs, which represent community-wide contact being relaxed from an 80% reduction in contact to 50%, plateau at approximately 34 daily infections when there is no “spike” in introduced cases and 30 infections in the “spike” setting. The brown graphs, which represent community-wide contact being relaxed from an 80% reduction in contact to 65%, average at approximately 15 infections without the “spike” and 16 infections with the “spike’. These slight differences are due to stochastic features in the model, where introduced infectious individuals are randomly allocated into modelled households.

The key difference between scenario *a* and both *b* and *c* is the ability of the latter two settings to increase workplace activity, by significantly lowering workplace non-attendance (WN), and with little effect on the infection rate. Settings *b* and *c* have higher community-wide contact reductions, 50% a nd 65% respectively, compared to *a* at 40%.

## Discussion

The timing of activation of social distancing measures has been a challenge facing public health authorities, and the rapid growth in cases and deaths in certain countries during the first three months of the COVID-19 pandemic highlights the criticality of early and robust responses, as has been analysed in a prior modelling study by the authors.^4^ A further challenge is knowing when to initiate relaxation of strong social distancing measures once ongoing COVID-19 virus transmission has declined, to avoid a rebound in case numbers. This study evaluated the easing of strong social distancing measures under two distinct settings: 1) for countries with high transmission rates arising from delays in activating response measures, as occurred in a number of European countries (i.e. Italy, Spain and the United Kingdom) and in New York State, USA and, 2) for countries which were able to rapidly respond to the initial ourbreak in China, by closing borders, activating robust social distancing measures, had capacity to rapidly diagnose cases, trace contacts and quarantine them. Countries such as Australia were thus able to contain the initial growth in case numbers and in some states, such as Western Australia, be close to virus elimination.^29^

Deciding on the timing and reduction in strength of social distancing measures as they are relaxed, to ensure that case numbers have declined to levels where health system resources are sustainable, is a challenge for governments. There is a need to balance what may be necessary to reduce the daily infection rate and take pressure off health care resources, with what a population can sustain, such as a long duration of highly restrictive measures, and the consequential negative impact this has on economic activity.

Modelling studies,^4,7,8^ and direct observation,^5,9^ have previously shown that robust and timely social distancing has the most effect in containing the spread of the COVID-19 virus. This study provides guidance as to the timing of relaxation of robust social distancing measures in both high and low transmission settings, for the cities of New York, USA and Perth, Australia respectively. They provide quantification of the effect on case numbers under alternative timings of social distancing easing, and how case numbers may be contained under a phased introduction of progressively weaker social distancing measures. The results demonstrate how to prevent a rebound in case numbers by adjusting the timing of relaxation and the magnitude (*i.e*. the strength) of reduced social distancing once the epidemic peak has passed.

As discussed by Di Domenico and colleagues,^6^ lifting “lockdown” without an informed exit strategy in place risks a second wave in infections and its impact on healthcare resources and deaths. Ending robust social distancing measures prematurely risks overwhelming health care resources, as is also suggested by our study.

In New York City social distancing measures were strengthened on 12^th^ April 2020 after those activated three weeks earlier failed to produce a significant decline in daily case numbers. If these measures were partially relaxed after six weeks our modelling suggests that the total number of infections occurring over the following three months will be approximately one million which, under asymptomatic and diagnosis percentage assumptions described in Table 1, may translate into a total of ~330,000 diagnosed cases. If these strict measures were held for an additional eight weeks before being partially relaxed, our modelling suggests that ~115,000 diagnosed cases would occur over that 100 day period. If strong social distancing was not relaxed we estimate that the number of diagnosed cases over the 100 day period would be ~100,000.

For a city the size of New York with an ~8·4 million population, the above estimated case numbers under alternative durations of robust social distancing give guidance as to the impact on health services when social distancing measures are eased. For example, the very flat epidemic curves which result from relaxing social distancing between eight and ten weeks after introduction (pink and green curves of Figure 2) have estimated diagnosed case peaks of approximately 2,500 per day. City-specific data relating diagnosed case numbers to hospitalization rates may then be used to estimate ventilator and ICU bed demand. While this will be higher than if the robust social distancing measures were not relaxed, if such estimated health care resource needs are within the capacity of the health care system then the decision to relax interventions becomes a tradeoff between managing infection numbers and increasing economic activity. When transmission has significantly reduced, relaxation of social distancing the allows schools and workplaces to safely reopen and community-wide activity to partly resume, with a resulting increase in economic activity.

In Australia the situation is quite different to that in the USA, the UK, France, and Italy, for example. Early interventions, including closing international borders and activating rigorous social distancing, has resulted in very low rates of transmission, with some states such as Western Australia tending to virus elimination, as of 25^th^ April 2020.^29^

Our model-based analyses indicate how social distancing measures may be successfully eased in a manner designed to increasingly restart the local economy while managing any significant increase in transmission. Such an increase in cases may occur from unseen spread among asymptomatic individuals or via movement of infected individuals due to the partial reopening of borders.

Managing the reopening of borders is a challenge to be faced by health authorities once ongoing virus transmission has reached low levels. Our analyses show how robust Australian social distancing measures have prevented arrival of introduced cases developing into a rapidly growing outbreak. Furthermore, a previous modelling analysis by the authors demonstrated how a significant rebound in daily cases may be mitigated by use of a robust, combined social distancing response.^4^

The COVID-19 transmission characteristics assumed in our model produced an unmitigated infection rate of 66%, which includes both symptomatic and asymptomatic cases, and a basic reproduction number of 2·2. Higher or lower R_0_ settings will affect case numbers under all the social distancing intervention considered. In the absence of definitive data on the proportion of infections which are asymptomatice we assumed a 35% asymptomatic proportion. Similarly, if the infectious period is longer than the three days assumed, the same caveat applies. However, sensitivity analyses indicate that changing the underlying model parameters to reflect these modifications does not affect the relative effectiveness of the social distancing measures.

The focus of this study is on measures to safely manage virus transmission within communities, whether towns or large cities. Halting movement between cities and countries is an additional measure to adopt in any pandemic situation. Others have examined these control measures for COVID-19 and pandemic influenza settings.^31,32^

## Data Availability

All data used is available in the public domain

## Contributors

GJM conceived the study. GJM and SX designed the study and collected data. SX and DP implemented the model and conducted experiments. GJM, SX and DP interpretated the results. GJM and SX drafted the Article. All authors contributed to the writing of the final version of the Article.

## Acknowledgement

This study was funded in part by the Department of Health, Government of Western Australia.

## Notes

### Competing Interest Statement

The authors have declared no competing interest.

### Funding Statement

Part funding from the Department of Health, Government of Western Australia is acknowledged.

